# AT(N) Framework in Older Adults with Epilepsy: Plasma Biomarkers and Associations with Demographic, Clinical, and Cognitive Features

**DOI:** 10.64898/2026.04.24.26351489

**Authors:** Kayela Arrotta, McKenna Williams, Nicolas R. Thompson, Katherine J. Bangen, Anny Reyes, Ifrah Zawar, Vineet Punia, Irene Wang, Jerry J. Shih, Lynn M. Bekris, Lisa Ferguson, Dace N. Almane, Jana E. Jones, Bruce P. Hermann, Robyn M. Busch, Carrie R. McDonald

## Abstract

**Background and Objectives:** Older adults with epilepsy have a 2- to 4-fold increased risk of dementia, including Alzheimer’s disease (AD), yet underlying mechanisms remain poorly defined. The NIA-AA classifies AD using amyloid (A), tau (T), and neurodegeneration [(N)] biomarkers. We applied this framework to characterize AT(N) profiles and clinical correlates in epilepsy.

**Methods:** Eighty-four older adults with focal epilepsy (mean age=66.3 years) from the Brain Aging and Cognition in Epilepsy (BrACE) study were classified as A+, T+, and/or (N)+ using plasma β-amyloid (Aβ) 42/40 ratio, phosphorylated tau 181 (p-tau181), and neurofilament light chain (NfL) levels, and grouped into normal, AD-continuum, and non-AD pathologic change. Demographic, clinical, and cognitive characteristics were compared. Cognition was assessed using the International Classification of Cognitive Disorders in Epilepsy (IC-CoDE) and the Montreal Cognitive Assessment (MoCA). Memory was examined using IC-CoDE memory domain classification, with word-list delayed recall analyzed separately. Associations with cognition were modeled using logistic and linear regression. Secondary analyses examined biomarkers continuously, including p-tau217, and substituted hippocampal volume for NfL.

**Results:** Only 32% of participants had normal biomarkers, while 37% were on the AD-continuum and 31% showed non-AD pathologic change. Participants with normal biomarkers were younger with shorter epilepsy duration, whereas *APOE*-ε4 carriers were enriched in the AD-continuum group. Early-onset compared to late-onset epilepsy (cutoff:≥55 years) showed higher odds of biomarker abnormality (aOR=8.84, 95% CI [2.35, 41.89], *P*=0.003), driven by elevated p-tau217, NfL, and greater amyloid burden. While categorical AT(N) profiles were not associated with cognition, higher p-tau181 levels were independently associated with lower word-list delayed recall (95% CI [−10.31, −0.86], *P*=0.021). Substituting hippocampal volume for NfL shifted more participants to normal profiles (48% vs. 32%) and fewer to non-AD pathologic change (15% vs. 31%).

**Discussion:** AT(N) biomarker profiles showed substantial heterogeneity, with higher abnormality rates than in aging populations, particularly among those with early-onset epilepsy. Continuous p-tau181 was associated with memory performance while categorical AT(N) profiles were not, and NfL and hippocampal volume showed discordant classifications, highlighting divergence across neurodegeneration markers. These findings underscore the complexity of applying AD-centric frameworks to epilepsy and support multimodal, epilepsy-adapted biomarker approaches to characterize neurodegenerative risk.

## Introduction

Older adults with epilepsy are at elevated risk for cognitive decline and dementia, including Alzheimer’s disease (AD), with epidemiologic studies suggesting an approximately 2–4-fold increase in dementia risk compared to the general population^1,2^. In parallel, seizures and epileptiform activity are increasingly recognized within the clinical spectrum of AD, and may both reflect and potentiate AD-related pathophysiology, supporting a bidirectional relationship between epileptic activity and neurodegenerative processes^3^. Despite this growing recognition, the biological mechanisms underlying the association between AD-related pathology and epilepsy remain incompletely understood.

The National Institute on Aging and Alzheimer’s Association (NIA-AA) created a research framework to define the underlying pathologic processes in AD and establish objective criteria for the diagnosis and staging of AD^4,5^. This research framework focuses on three primary biomarkers linked to AD: amyloid (A), tau (T), and neurodegeneration [(N)]. A key strength of the AT(N) framework is that it distinguishes amyloid, tau, and neurodegeneration as separable biological processes, enabling examination of their independent and combined contributions to disease stage and clinical expression, rather than treating AD-related pathology as a unitary construct. This approach has advanced our understanding of the heterogeneous pathways to cognitive decline in broader aging populations and has informed the development of targeted interventions^6,7^. With the emergence of blood-based biomarkers, the AT(N) framework can now be applied more broadly, enabling scalable and minimally invasive characterization of neurodegenerative pathology^8^.

Emerging evidence indicates that AD-related biomarkers are also relevant to epilepsy, supporting shared mechanisms linking epileptic activity and neurodegeneration^9,10^. Application of the AT(N) framework to epilepsy could therefore provide a systematic approach to characterizing pathological burden and understanding how amyloid, tau, and neurodegeneration, individually and in combination, contribute to seizure characteristics and cognitive difficulties commonly experienced by older adults with epilepsy. Indeed, each component of the AT(N) framework has been independently linked to epilepsy pathophysiology and cognition.

β-amyloid (Aβ) is related to neurodegeneration and has been associated with epileptogenesis,^11,12^ particularly in late-onset unexplained epilepsy (LOUE)^11,13^. Evidence also suggests that amyloid burden may affect cognitive status in epilepsy. Specifically, Aβ accumulation has been associated with greater learning and executive functioning deficits in individuals with drug-resistant epilepsy^14,15^, and poorer global cognitive scores in LOUE^16,17^.

Similarly, aggregated tau (T) can increase neurodegeneration and neuronal hyperexcitability, and may play a crucial role in epileptogenesis. Most studies have found increased tau levels in epilepsy^18–20^, and animal models have demonstrated that tau may be a useful prognostic and disease-progression biomarker in epilepsy^21,22^. In temporal lobe epilepsy, elevated hyperphosphorylated tau is associated with worse seizure severity and frequency, status epilepticus (SE), and may contribute to cognitive impairments, specifically in verbal learning and attention^15,23–25^.

Neurofilament light chain (NfL), a marker of neuronal injury, is a biomarker recognized within the AT(N) framework to measure neurodegeneration and has been connected to a number of neurodegenerative conditions, including AD, frontotemporal dementia, and dementia with Lewy bodies^26,27^. The evidence on the association between NfL and epilepsy is mixed, with one study showing significantly increased NfL in older adults with epilepsy compared to controls^28^, while other studies have not found significant differences in persons with epilepsy^3^. However, emerging evidence suggests potential prognostic value with higher levels of NfL associated with SE^29^ and correlation with global cognitive function in epilepsy populations^9^.

Together, these findings suggest that components of the AT(N) framework may capture biologically meaningful variation in epilepsy, although their combined application remains underexamined. Findings from a pilot study on LOUE revealed that cerebrospinal fluid (CSF) AT(N) profiles were similar across LOUE individuals with and without cognitive impairment, while non-amyloid related processes seemed more contributory in non-epilepsy controls with cognitive impairment^17^. However, this study had a limited sample size and focused only on those with LOUE. Additional research is needed to clarify whether AT(N) classifications in epilepsy reflect AD-related pathology, epilepsy-specific processes, or both, and to better characterize the pathological basis of cognitive impairment and dementia risk in this population.

In this study, we applied the AT(N) framework using plasma biomarkers in a well-characterized cohort of older adults with epilepsy, including both early-onset epilepsy and LOUE. We aimed to (1) characterize the distribution of AT(N) biomarker profiles and (2) examine their associations with demographic, clinical, and cognitive variables. We further evaluated biomarkers continuously and explored alternative neurodegeneration markers to better understand how biomarker selection influences classification and clinical interpretation in epilepsy.

## Materials and Methods

### Study Design

Brain Aging and Cognition in Epilepsy (BrACE) is a prospective, longitudinal multicenter study focused on adults ≥55 years of age with early-onset (epilepsy-onset age <55 years) or LOUE (epilepsy-onset age ≥55 years).

### Standard Protocol Approvals, Registrations, and Patient Consents

The study was approved by the Institutional Review Boards at the University of California, San Diego (UCSD; approval #210422), the Cleveland Clinic (CC; approval #22-079), and the University of Wisconsin-Madison (UWM; approval #2021-0929). Written informed consent was obtained from all participants prior to enrollment.

### Participants

Participants were aged ≥55 years old with an established diagnosis of epilepsy confirmed by a board-certified epileptologist according to International League Against Epilepsy criteria^30^.

Participants were included if they had no prior epilepsy-related neurosurgery and were proficient in English for completion of the neuropsychological assessment. Participants were excluded if they had a history of clinical stroke, a space-occupying lesion on MRI, or a diagnosis of primary psychosis, dementia, or other neurological diseases other than epilepsy at baseline.

### Measures

*Demographic and Seizure Characteristics.* Demographic, clinical data, and seizure-related information were obtained for participants using standardized data collection forms across study sites and electronic medical records. Demographic variables included age, sex, years of education, race, ethnicity, and handedness.

Seizure-related variables included age at epilepsy onset, duration of epilepsy, side of epilepsy (left, right, bilateral, or unknown), site of epilepsy (frontal, temporal, frontotemporal, or unknown), seizure frequency, days since last seizure (relative to blood draw), history of SE, history of focal to bilateral tonic-clonic seizures, drug-resistance status, and number of anti-seizure medications (ASMs).

Clinical factors relevant to biomarker interpretation, including chronic kidney disease (CKD), family history of Alzheimer’s disease (AD), and APOE status, were also recorded.

*Biomarkers and AT(N) Profiles.* Aβ40, Aβ42, and total-tau were measured using the Neurology 3-Plex A Advantage Kit, and phosphorylated tau 181 (p-tau181) was measured using the p-tau181 Advantage V2.1 Assay Kit (single-plex), on a single molecule array platform (Simoa SR-X platform, 101995 and 104111, Quanterix; Billerica, MA). NfL was measured using the R-PLEX Human Neurofilament L Assay from Meso Scale Discovery (K1517XR, MSD; Rockville, MD). P-tau217 was measured using the S-PLEX Human Tau (pT217) Kit from Meso Scale Discovery (K151APFS, MSD; Rockville, MD). All assays were processed and performed according to the manufacturer’s instructions. Plasma biomarker cutpoints were derived from CSF thresholds previously validated against PET using an amyloid positivity threshold of ≥24 Centiloids^31^.

Participants were classified as A+, T+, and/or (N)+ based on plasma Aβ42/Aβ40 pg/mL ratio ≤ 0.041619939, p-tau181 pg/mL ≥ 25.21083668, and NfL pg/mL ≥ 84.58311893, respectively. Due to limited sample size across the eight possible biomarker combinations, profiles were grouped into three categories: 1) normal biomarkers (A-T-N-), 2) AD continuum (A+T-N-, A+T+N-, A+T-N+, A+T+N+), and 3) non-AD pathologic change (A-T+N-, A-T-N+, A-T+N+). Given emerging evidence that p-tau217 may be a particularly sensitive biomarker for both amyloid and tau pathology^32^, this biomarker was additionally analyzed as a continuous measure, as a validated cutoff was not available at the time of the study.

*Neuropsychological Data.* Participants completed a comprehensive neuropsychological evaluation assessing memory, language, attention/processing speed, and executive function. The International Classification of Cognitive Disorders in Epilepsy (IC-CoDE) was used to develop cognitive phenotypes^33^. Due to the absence of visuospatial measures in the battery, a four-domain classification schema was used. Participants were classified as having an impaired IC-CoDE phenotype if they demonstrated impairment in one or more cognitive domains as defined by two or more test scores within a domain falling below 1.0 standard deviation based on normative data (i.e., single domain, bi-domain, and generalized impaired phenotypes were combined). Participants were classified as having an intact IC-CoDE phenotype if no cognitive domains were impaired. The Montreal Cognitive Assessment (MoCA) was also used as a continuous measure of global cognition, which has demonstrated validity in this epilepsy cohort^34^. Given the relevance of memory in epilepsy and AD biomarker research, memory was evaluated both categorically (IC-CoDE memory domain impairment), and continuously, using the delayed recall T-scores from the Rey Auditory Verbal Learning Test (RAVLT), a word-list recall measure sensitive to verbal memory dysfunction in epilepsy^35^.

*Hippocampal Volume.* Although NfL is one of the most commonly used blood-based biomarkers for neurodegeneration, elevated NfL levels in epilepsy populations may reflect seizure-related neuronal injury rather than neurodegenerative processes^36^. Given this potential confound, we examined hippocampal volume as an alternative marker for neurodegeneration^37^. Hippocampal volume was obtained from T1-weighted images acquired at each site on a 3T MRI system (UCSD = GE Discovery MR750 SPGR, voxel size =1mm isotropic, TR=0.006, TE=0.002, TI=1.06, flip angle=8; CC = Siemens Prisma MPRAGE, voxel size = 1mm isotropic, TR=2.3, TE=0.002, TI=.9, flip angle=9; UWM = GE Signa Premier, SPGR, TR=0.007, TE=0.002, TI=.4, flip angle = 11) and segmented using automated volumetric segmentation software (Neuroquant software, Cortech, Inc). Normative percentiles for hippocampal volumes were adjusted for age, sex, and intracranial volume provided by the Neuroquant software package.

Total hippocampal volume was missing for six participants who did not undergo MRI, while ipsilateral hippocampal volume was missing for an additional 13 participants due to unknown seizure lateralization. Given the more complete data availability, total hippocampal volume (sum between left and right volumes) was used in analyses. As there are no standard guidelines or cutpoints^37^, two cutpoint values, below 10^th^ and 25^th^ percentiles, were used to define neurodegeneration based on total hippocampal volume.

### Statistical Analyses

AT(N) profiles were created for all participants with complete biomarker data, and the frequency and percentage of each profile was summarized. Descriptive statistics of participants and clinical characteristics were computed, stratified by the three-category AT(N) profiles. Means with standard deviations (when normality assumptions were met) or medians with interquartile ranges (when normality assumptions were violated) summarized continuous variables, and frequency with percentages summarized categorical variables. Comparisons were made using one-way analysis of variance (ANOVA) or Kruskal-Wallis tests for continuous variables and Fisher’s exact test for categorical variables. Where initial group comparisons revealed significant differences, post-hoc analyses were conducted to further characterize associations between epilepsy characteristics and individual biomarkers.

To examine hippocampal volume as an alternative neurodegeneration marker, AT(N) profiles were refit substituting total hippocampal volume percentile for NfL, and demographic and clinical characteristics of the resulting profiles were compared using the same procedures described above. The relationship between NfL and continuous total hippocampal volume was examined using Spearman’s rank correlation, given mild non-normality of the hippocampal volume distribution. All analyses including hippocampal volume percentile were additionally rerun using ipsilateral hippocampal volume as a sensitivity analysis.

The association between AT(N) profiles and IC-CoDE was modeled using logistic regression, where IC-CoDE phenotype (impaired vs. intact) and IC-CoDE memory domain (impaired vs. intact) were the dependent variables, and AT(N) profile group (normal biomarkers, AD continuum, and non-AD pathologic change) was the independent variable. Firth’s penalized likelihood method was used for all logistic regression models with binary outcomes (IC-CoDE phenotype and memory domain, intact vs. impaired). Linear regression was used to model the associations between AT(N) profiles and MoCA total score, and RAVLT delayed recall T-score, separately.

We also fit separate models where biomarkers were analyzed as continuous predictors (Aβ42/Aβ40, p-tau181, NfL, p-tau217, and hippocampal volume percentile). The p-tau181 variable was log-transformed to address non-normality, and sensitivity analyses were performed excluding one outlier. Bootstrapped confidence intervals for hippocampal volume percentile were also employed for these models as a sensitivity analysis to address distributional concerns.

For each of these models, both unadjusted (i.e., no covariates) and covariate-adjusted models were fit. In covariate-adjusted models, the following covariates, chosen a priori, were included: age, years of education, age at epilepsy onset, side of epilepsy (left vs. right/bilateral/unknown), site of epilepsy (temporal vs. frontotemporal/frontal/NOS), and presence of CKD, given known effects of renal function on blood biomarker concentrations^38^. For the models that examined Aβ42/Aβ40, p-tau181, NfL, and p-tau217 continuously, we additionally adjusted for storage time.

All analyses were performed in R, version 4.4.1. All tests were two-sided, and p-values < 0.05 were considered statistically significant.

### Data Availability

The data that support the findings of this study are available from the corresponding author, upon reasonable request.

## Results

### Study Sample

A total of 93 participants with epilepsy were included, of whom nine were excluded due to missing AT(N) data, resulting in a final sample of 84 participants. Participant characteristics are summarized in Table 1.

**Table 1.** Demographic, seizure, cognitive, and medical characteristics stratified by AT(N) profile.

|  | Total<br>(n = 84) | Normal<br>Biomarkers<br>(n = 27) | AD Continuum<br>(n = 31) | Non-AD<br>Pathologic<br>Change<br>(n = 26) | P |
| --- | --- | --- | --- | --- | --- |
| **Demographics** |  |  |  |  |  |
| Age, years, mean (SD) | 66.3 (6.9) | 63.7 (5.5) | 68.0 (7.8) | 67.0 (6.5) | **.045** |
| Female sex, n (%) | 48 (57.1%) | 15 (55.6%) | 18 (58.1%) | 15 (57.7%) | >.999 |
| Hispanic ethnicity, n (%) | 6 (7.1%) | 1 (3.7%) | 2 (6.5%) | 3 (11.5%) | .585 |
| White race, n (%) | 72 (85.7%) | 23 (85.2%) | 26 (83.9%) | 23 (88.5%) | .997 |
| Years of education, mean (SD) | 15.0 (2.6) | 15.1 (2.6) | 15.0 (2.7) | 14.8 (2.4) | .868 |
| **Seizure Characteristics** |  |  |  |  |  |
| Age at epilepsy onset, years, mean (SD) | 44.0 (20.5) | 49.3 (16.1) | 42.2 (20.6) | 40.7 (23.8) | .259 |
| Epilepsy duration, years, median (IQR) | 20 (5, 37.5) | 11 (3, 20.5) | 21 (11, 43) | 22 (5, 41.5) | **.027** |
| Seizure frequency, n (%) |  |  |  |  | .236 |
| Innumerable to monthly | 12 (14.3%) | 6 (22.2%) | 5 (16.1%) | 1 (3.8%) |  |
| At least yearly | 33 (39.3%) | 12 (44.4%) | 10 (32.3%) | 11 (42.3%) |  |
| Unknown | 39 (46.4%) | 9 (33.3%) | 16 (51.6%) | 14 (53.8%) |  |
| FTBTC history, Yes, n (%) | 28 (33.3%) | 9 (33.3%) | 12 (38.7%) | 7 (26.9%) | .467 |
| SE history, Yes, n (%) | 2 (2.4%) | 0 (0.0%) | 1 (3.2%) | 1 (3.8%) | .699 |
| Drug-resistance status, n (%) |  |  |  |  | .303 |
| Drug-responsive | 27 (32.1%) | 5 (18.5%) | 11 (35.5%) | 11 (42.3%) |  |
| Drug-resistant | 52 (61.9%) | 19 (70.4%) | 19 (61.3%) | 14 (53.8%) |  |
| Unknown | 5 (6.0%) | 3 (11.1%) | 1 (3.2%) | 1 (3.8%) |  |
| Side of epilepsy, n (%) |  |  |  |  | .926 |
| Right | 19 (22.6%) | 6 (22.2%) | 6 (19.4%) | 7 (26.9%) |  |
| Left | 35 (41.7%) | 11 (40.7%) | 15 (48.4%) | 9 (34.6%) |  |
| Bilateral | 15 (17.9%) | 4 (14.8%) | 6 (19.4%) | 5 (19.2%) |  |
| Unknown | 15 (17.9%) | 6 (22.2%) | 4 (12.9%) | 5 (19.2%) |  |
| Site of epilepsy, n (%) |  |  |  |  | .439 |
| Temporal | 41 (48.8%) | 10 (37.0%) | 15 (48.4%) | 16 (61.5%) |  |
| Frontal | 4 (4.8%) | 3 (11.1%) | 1 (3.2%) | 0 (0.0%) |  |
| Frontotemporal | 20 (23.8%) | 6 (22.2%) | 8 (25.8%) | 6 (23.1%) |  |
| Unknown | 19 (22.6%) | 8 (29.6%) | 7 (22.6%) | 4 (15.4%) |  |
| Number of ASMs, median (IQR) | 1 (1, 2) | 1 (1, 2) | 1 (1, 2) | 2 (1, 2) | .679 |
| **Cognition** |  |  |  |  |  |
| MoCA total score, mean (SD) | 24.6 (3.0) | 25.5 (2.5) | 24.1 (3.0) | 24.4 (3.4) | .175 |
| IC-CoDE phenotype impaired, n (%) | 27 (32.1%) | 9 (33.3%) | 7 (22.6%) | 11 (42.3%) | .274 |
| RAVLT delayed recall T-score, mean (SD) | 44.6 (10.7) | 46.3 (12.0) | 43.7 (10.5) | 43.8 (9.6) | .599 |
| Memory domain impaired, n (%) | 11 (13.1%) | 3 (11.1%) | 4 (12.9%) | 4 (15.4%) | .923 |
| **Other Medical** |  |  |  |  |  |
| Family AD history, Yes, n (%) | 15 (17.9%) | 6 (22.2%) | 6 (19.4%) | 3 (11.5%) | .450 |
| APOEε4 positive, Yes, n (%) | 19 (22.6%) | 5 (18.5%) | 11 (35.5%) | 3 (11.5%) | **.031** |
| CKD positive, n (%) | 4 (4.8%) | 0 (0.0%) | 1 (3.2%) | 3 (11.5%) | .151 |

**Note.** Continuous variables: mean (SD) or median (IQR). Categorical variables: n (%). Group comparisons: ANOVA or Kruskal-Wallis (continuous) and Fisher's exact test (categorical). Bold P-values indicate significance ( $P < .05$ ). Binary variables (FTBTC, SE, Family AD history, APOEε4) show affirmative response only; unknown/missing not shown. Race: other categories each <5% (Black 6.0%, Asian 1.2%, American Indian/Alaska Native 1.2%, Multiracial 3.6%, Unknown 2.4%). MoCA = Montreal Cognitive Assessment; RAVLT = Rey Auditory Verbal Learning Test; IC-CoDE = International Classification of Cognitive Disorders in Epilepsy; FTBTC = focal to bilateral tonic-clonic seizures; SE = status epilepticus; ASMs = anti-seizure medications; CKD = chronic kidney disease; AD = Alzheimer's disease; IQR = interquartile range.

The mean participant age was 66.3 years (standard deviation [SD] = 6.9), with an average of 15.0 years of education (SD = 2.6). The cohort was predominantly white (85.7%) and female (57.1%). The average age of epilepsy onset was 44.0 years (SD = 20.5), with a median epilepsy duration of 20 years (interquartile range [IQR] = 5, 37.5). Participants were usually taking one anti-seizure medication (IQR = 1,2). Most participants had temporal lobe epilepsy (48.8%), with left- sided seizure onset being the most common (41.7%). While 61.9% met criteria for drug-resistant epilepsy (DRE), only three participants experienced a seizure within 24 hours of the blood draw.

### AT(N) Profiles and Demographic and Seizure Characteristics

Of the 84 participants, 27 (32.1%) had normal blood-based AD biomarkers, 31 (36.9%) were classified within the AD continuum, and 26 (31.0%) demonstrated non-AD pathologic change (Figure 1). Demographic, seizure, and clinical characteristics stratified by AT(N) profile are presented in Table 1. Participants in the normal biomarker group were significantly younger and had significantly shorter epilepsy duration. Post-hoc analyses examining duration of epilepsy and individual biomarkers revealed that longer duration of epilepsy was associated with higher levels of NfL (per 10-year *B* = 5.72, 95% CI [1.26, 10.18], *P* = 0.013, partial η*²* = 0.096). No significant associations were found between epilepsy duration and other biomarkers (i.e., p-tau217, p-tau181 or Aβ42/Aβ40). Participants in the AD continuum group were more likely to carry one or two *APOE*-ε4 alleles. No other demographic or seizure-related variables differed significantly across AT(N) groups.

**Figure 1.**
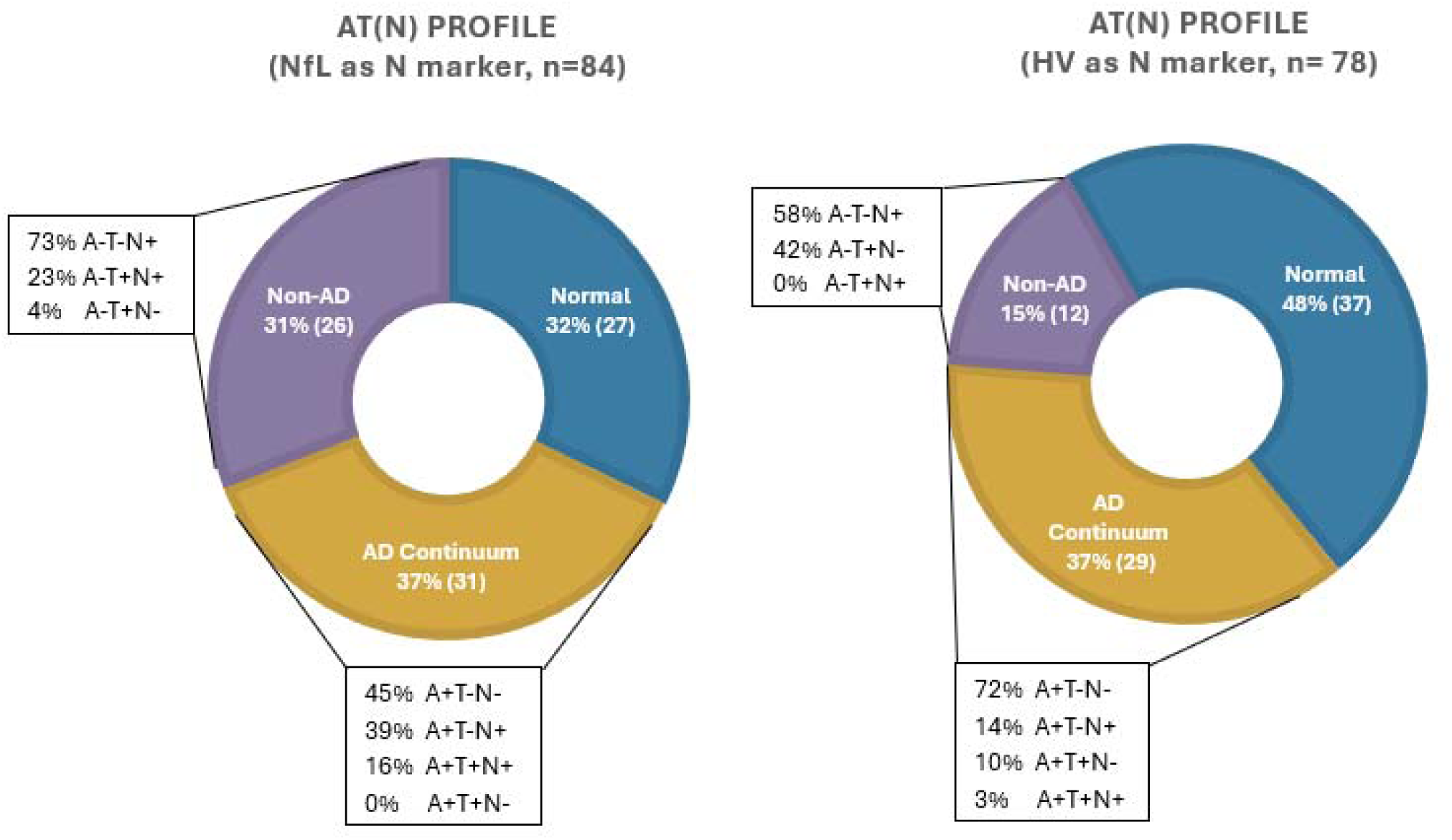
AT(N) profiles in older adults with epilepsy by NfL or hippocampal volume (HV) as N marker. Donut charts illustrating the distribution of AT(N) profiles: normal (A−T−N−), AD continuum (any A+ profile), and non-AD pathologic change (A−T+ and/or N+), when neurodegeneration (N) is indexed by neurofilament light chain (NfL; *n*=84; left) or total hippocampal volume (HV; *n*=78; right). Callout boxes display the constituent biomarker combinations within each profile group. Substitution of HV for NfL resulted in a higher proportion classified as normal (48% vs. 32%) and a lower proportion with non-AD pathologic change (15% vs. 31%), with AD continuum proportions remaining stable (37% in both). AT(N) = amyloid/tau/neurodegeneration framework; AD = Alzheimer’s disease; NfL = neurofilament light chain; HV = hippocampal volume

Although age of epilepsy onset was not significantly associated with blood-based AT(N) profile, post-hoc analyses examined AT(N) profiles in early-onset epilepsy (Mean = 30.4, SD = 14.6) versus LOUE (Mean = 64.1, SD = 6.4). There were no significant differences between the two groups. However, when AT(N) groups were collapsed into normal versus abnormal blood-based biomarker profiles, early-onset epilepsy was associated with higher odds of biomarker abnormality compared to LOUE (aOR = 8.84, 95% CI [2.35, 41.89], *P* = 0.003), after adjusting for age, years of education, side and site of epilepsy, and presence of CKD (Figure 2).

**Figure 2.**
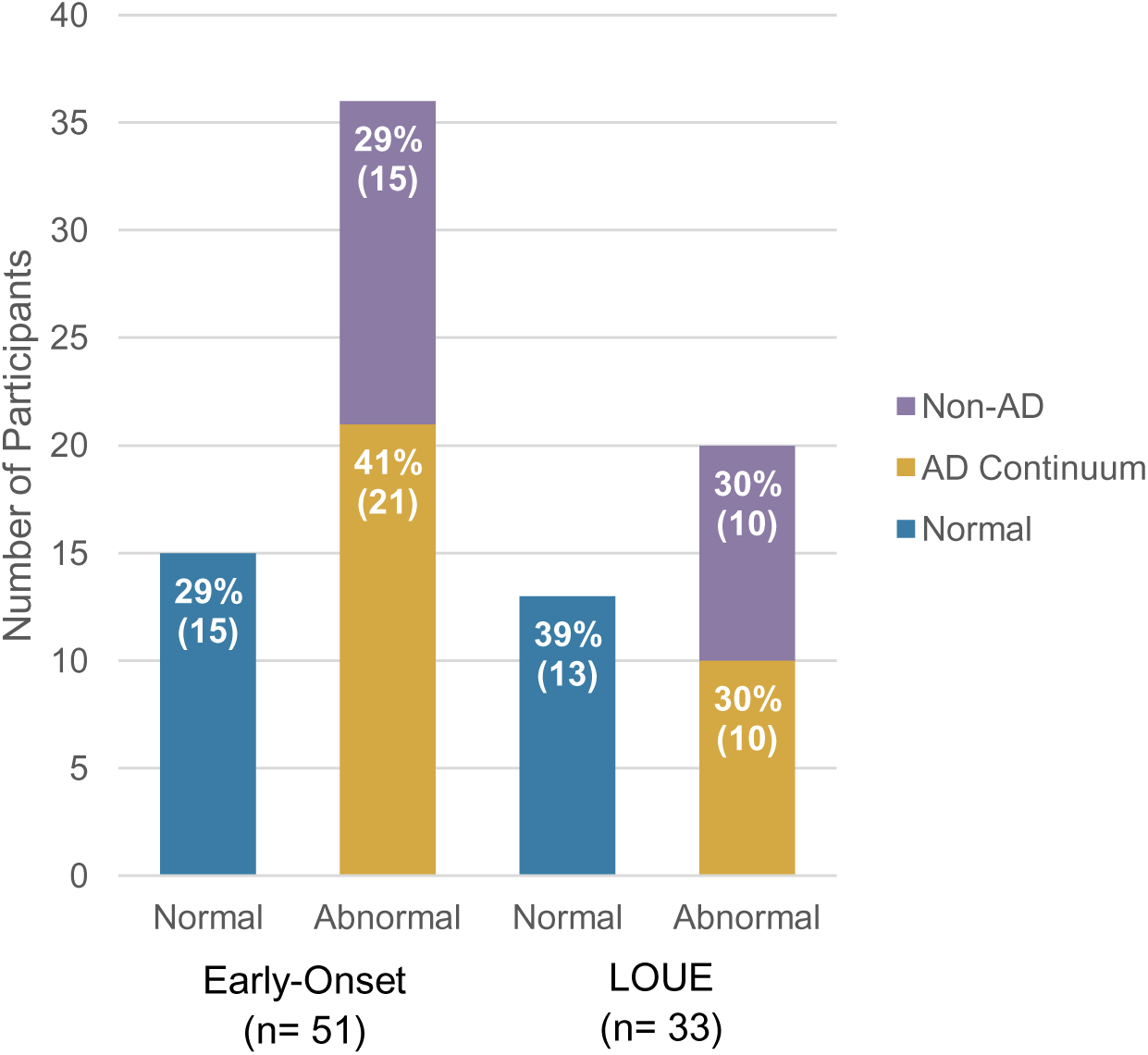
AT(N) profiles in early-onset vs LOUE. Stacked bar charts displaying the number and percentage of participants classified as normal, AD continuum, or non-AD pathologic change within normal and abnormal AT(N) profile categories, stratified by epilepsy onset group: early-onset (*n*=51) and late-onset unexplained epilepsy (LOUE; *n*=33). A higher proportion of early-onset participants showed any biomarker abnormality (71% vs. 61%), driven largely by a greater proportion classified in the AD continuum (41% vs. 30%). AT(N) = amyloid/tau/neurodegeneration framework; AD = Alzheimer’s disease; LOUE = late-onset unexplained epilepsy.

Examination of individual biomarkers revealed that the early-onset group had higher levels of NfL (*B* = 29.65, 95% CI [10.71, 48.59], *P* = 0.003, partial η*²* = 0.002) and p-tau217 (*B* = 1.30, 95% CI [0.33, 2.28], *P* = 0.010, partial η*²* = 0.008), as well as greater amyloid burden as reflected by a lower Aβ42/Aβ40 ratio (*B* = −0.004, 95% CI [-0.008, −0.00003], *P* = 0.048, partial η*²* = 0.031). No significant differences were observed for p-tau181 (Figure 3).

**Figure 3.**
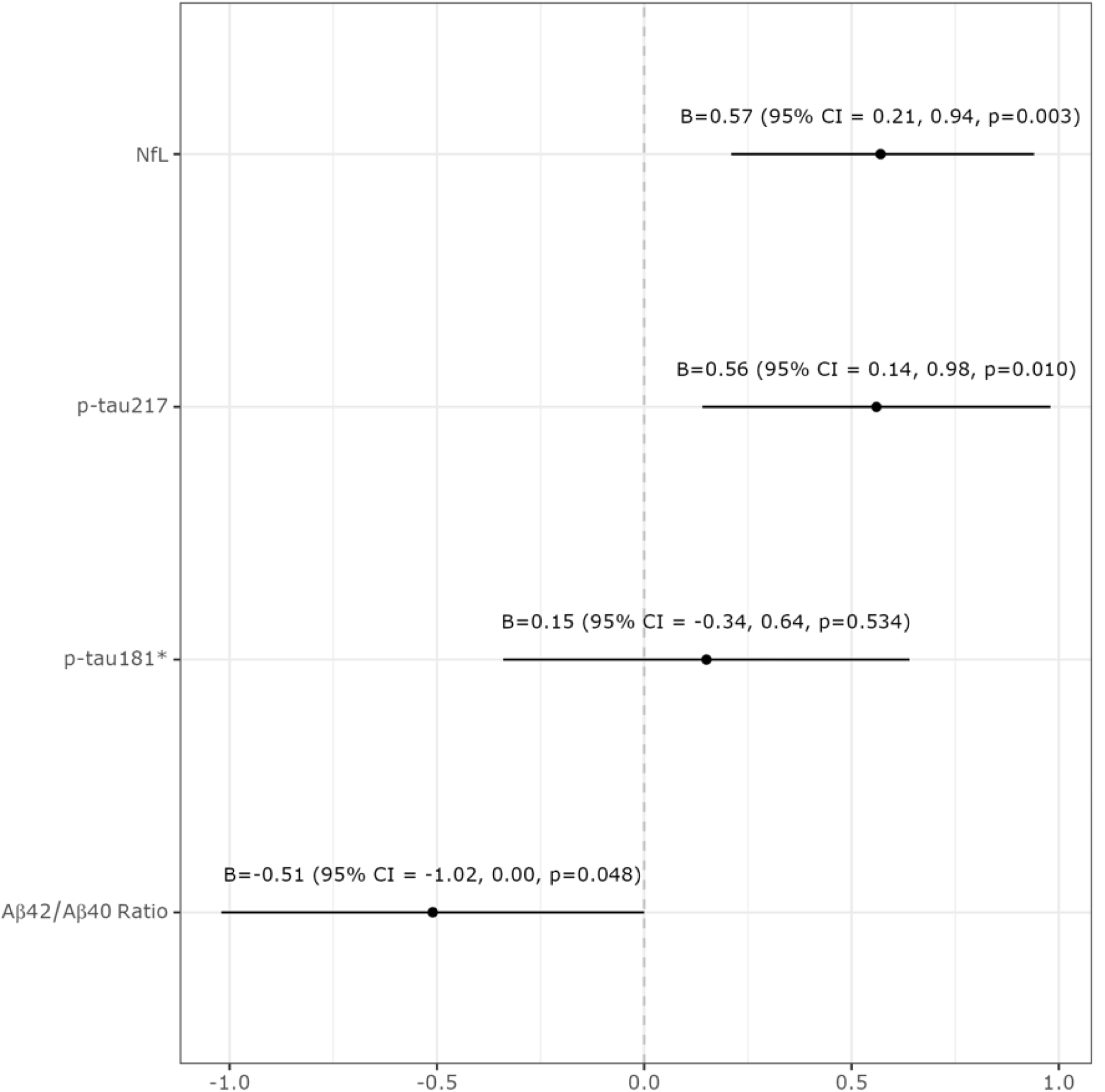
Individual plasma biomarkers in early-onset vs LOUE. Points represent standardized regression coefficients (B) and horizontal lines represent 95% confidence intervals. Positive values indicate higher biomarker levels in the early-onset group relative to LOUE. Early-onset epilepsy was associated with higher NfL (*B*=0.57, 95% CI [0.21, 0.94], *P*=0.003), higher p-tau217 (*B*=0.56, 95% CI [0.14, 0.98], *P*=0.010), and lower Aβ42/Aβ40 ratio (*B*=−0.51, 95% CI [−1.02, 0.00], *P*=0.048), but did not differ on p-tau181 (*B*=0.15, 95% CI [−0.34, 0.64], *P*=0.534). Models were adjusted for age, years of education, side of epilepsy focus, site of epilepsy focus, presence of chronic kidney disease, and storage time. *p-tau181 was log-transformed prior to analysis due to non-normal distribution. AT(N) = amyloid/tau/neurodegeneration framework; NfL = neurofilament light chain; LOUE = late-onset unexplained epilepsy; CI = confidence interval.

Figure 4 illustrates the distribution of participants across epilepsy onset group, seizure localization, DRE status, and AT(N) classification. Visual inspection suggests heterogeneous pathways to biomarker profiles, with early-onset epilepsy showing apparent enrichment in the AD continuum classification.

**Figure 4.**
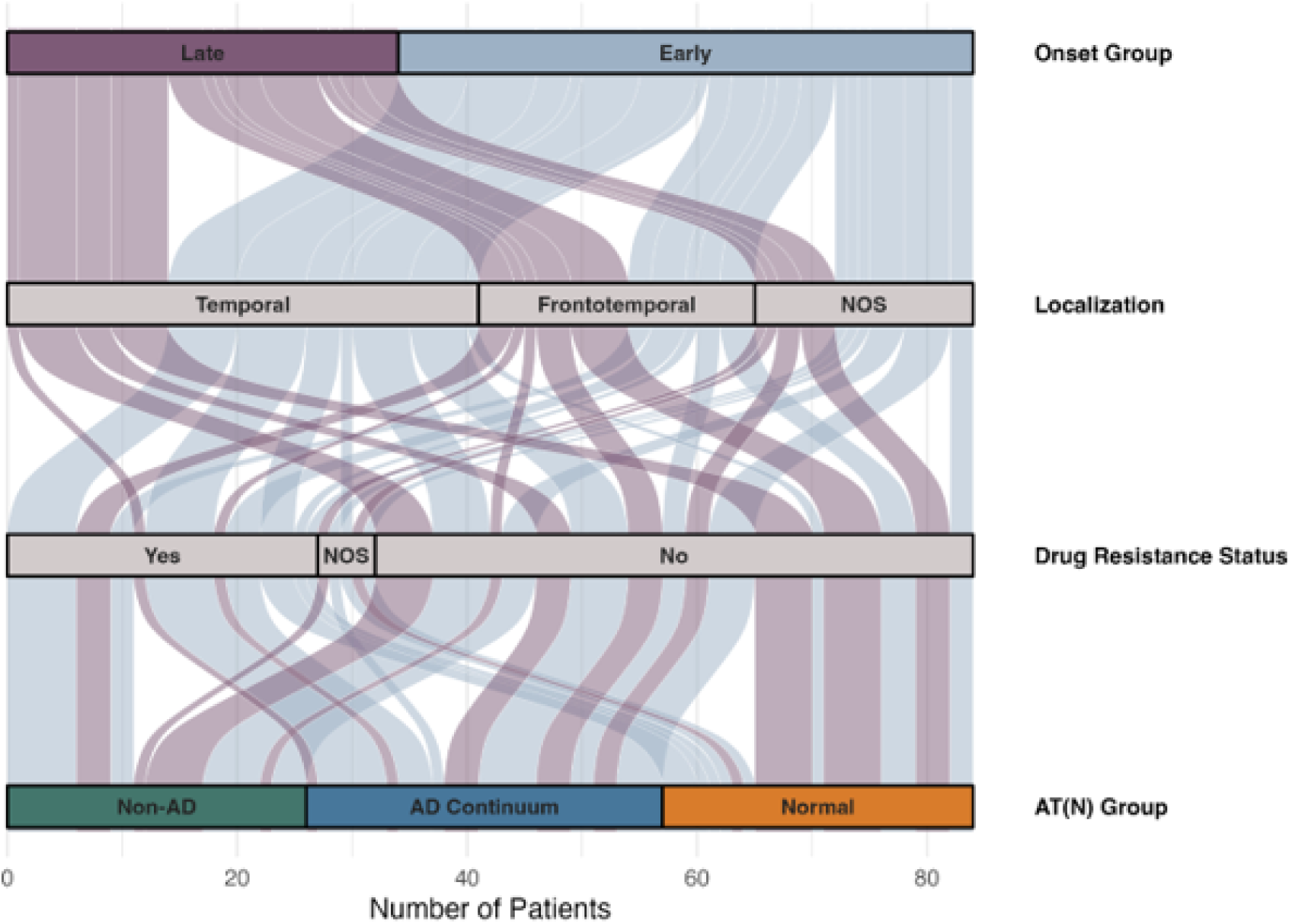
AT(N) classification stratified by onset, seizure localization, and drug-resistance status. Alluvial diagram illustrating the flow of participants (*n*=84) from epilepsy onset group (early-onset, late-onset) through seizure localization (temporal, frontotemporal, not otherwise specified [NOS]) and drug-resistance status (drug-resistant, NOS, drug-responsive) to final AT(N) biomarker classification (non-AD pathologic change, AD continuum, normal). Stream width is proportional to the number of participants. Notably, early-onset participants showed a disproportionate flow toward the AD continuum regardless of seizure localization or drug-resistance status, suggesting that epilepsy chronicity and duration, rather than focal characteristics or pharmacological response, may be more closely linked to AD-related biomarker burden. NOS = not otherwise specified; AT(N) = amyloid/tau/neurodegeneration framework; AD = Alzheimer’s disease.

### Hippocampal Volume as a Marker of Neurodegeneration

NfL and continuous total hippocampal volume were not significantly correlated (Spearman’s *r* = −0.155, *P* = 0.174). In general, fewer participants were positive for neurodegeneration using total hippocampal volume as compared to NfL. Using the more liberal cutoff of total hippocampal volume below the 25^th^ percentile, 37 participants (47.4%) were classified as having normal biomarker, 29 (37.2%) on the AD continuum, and 12 (15.4%) with non-AD pathologic change (Figure 1). Those who were classified as having non-AD pathologic change had a higher number of ASMs (*P* = 0.014), while no other demographic or clinical variables were associated with AT(N) profile when using hippocampal volume. Results did not significantly differ using the ipsilateral hippocampal volume.

### AT(N) Profiles and Cognition

None of the AT(N) profiles, continuous ATN variables, p-tau217, or hippocampal volume (bootstrapped models) were significantly associated with IC-CoDE phenotype or MoCA total score in unadjusted or covariate-adjusted models (Table 2). None of the covariates were associated with IC-CoDE phenotype, while more years of education was associated with higher MoCA total score.

**Table 2.** AT(N) and individual biomarker associations with cognitive outcomes.

| Predictor | MoCA Total |  | RAVLT Delayed Recall |  | IC-CoDE Binary |  | IC-CoDE Memory Domain |  |
| --- | --- | --- | --- | --- | --- | --- | --- | --- |
|  | B (95% CI) | P | B (95% CI) | P | OR (95% CI) | P | OR (95% CI) | P |
| **Categorical: Three-Category AT(N) (NfL as N marker)** |  |  |  |  |  |  |  |  |
| AD Continuum (vs. Normal) | -1.18<br>(-2.80, 0.45) | .153 | -4.41<br>(-10.58, 1.76) | .158 | 0.51<br>(0.13, 1.84) | .308 | 1.35<br>(0.27, 7.03) | .711 |
| Non-AD Change (vs. Normal) | -0.68<br>(-2.38, 1.03) | .432 | -3.86<br>(-10.33, 2.61) | .238 | 1.20<br>(0.33, 4.34) | .784 | 1.61<br>(0.32, 8.71) | .558 |
| **Categorical: Three-Category AT(N) (Hippocampal Volume as N, 25th Percentile)** |  |  |  |  |  |  |  |  |
| AD Continuum (vs. Normal) | -1.31<br>(-2.73, 0.11) | .070 | -2.64<br>(-8.16, 2.87) | .343 | 0.59<br>(0.18, 1.86) | .373 | 1.33<br>(0.32, 5.60) | .691 |
| Non-AD Change (vs. Normal) | -0.84<br>(-2.76, 1.09) | .389 | -0.42<br>(-7.94, 7.11) | .913 | 1.55<br>(0.33, 7.14) | .569 | 1.97<br>(0.30, 10.95) | .452 |
| **Continuous Biomarkers** |  |  |  |  |  |  |  |  |
| Aβ42/Aβ40 | 0.79<br>(-0.02, 1.60) | .055 | 0.81<br>(-2.37, 4.00) | .613 | 1.60<br>(0.80, 3.50) | .207 | 1.19<br>(0.54, 3.04) | .685 |
| P-tau181<sup>a</sup> | -1.06<br>(-2.32, 0.20) | .097 | -5.59<br>(-10.31, -0.86) | **.021** | 2.29<br>(0.85, 7.83) | .129 | 2.01<br>(0.73, 6.05) | .163 |
| P-tau181<sup>a</sup><br>(Outlier Removed) | -1.19<br>(-2.84, 0.47) | .157 | -7.58<br>(-13.77, -1.39) | **.017** | 1.74<br>(0.46, 7.02) | .419 | 2.01<br>(0.73, 6.05) | .163 |
| NfL | -0.04<br>(-0.21, 0.13) | .669 | -0.25<br>(-0.91, 0.41) | .453 | 1.03<br>(0.90, 1.17) | .711 | 1.04<br>(0.88, 1.23) | .610 |
| P-tau217 | -0.09<br>(-0.42, 0.24) | .583 | -0.05<br>(-1.28, 1.19) | .937 | 1.07<br>(0.82, 1.37) | .617 | 0.98<br>(0.70, 1.30) | .889 |
| HV Percentile<sup>b</sup> | -0.07 (-0.36, 0.17) | .611 | -0.52 (-1.65, 0.49) | .352 | 1.26 (1.03, 1.60) | .054 | 1.05 (0.83, 1.34) | .710 |

*Note.* All models adjusted for age, years of education, age of epilepsy onset, side of epilepsy focus, site of epilepsy focus, and presence of chronic kidney disease. Models examining continuous biomarkers individually were additionally adjusted for their respective storage time. *B* = unstandardized coefficient; OR = odds ratio; CI = confidence interval; NfL = neurofilament light chain; HV = hippocampal volume; MoCA = Montreal Cognitive Assessment; RAVLT = Rey Auditory Verbal Learning Test; IC-CoDE = International Classification of Cognitive Disorders in Epilepsy. <sup>a</sup>Log-transformed due to non-normal distribution. <sup>b</sup>Confidence intervals and p-values for HV Percentile reflect bootstrapped estimates (1,000 samples) to address mild non-normality of hippocampal volume distribution. Bold indicates $P < 0.05$ .

No significant associations were observed between IC-CoDE memory domain impairment and AT(N) profiles, continuous ATN variables, p-tau217, or hippocampal volume (bootstrapped models) in unadjusted or covariate-adjusted models. None of the independent variables of interest were associated with word-list delayed recall score in unadjusted models. In covariate-adjusted models, log-transformed p-tau181 was associated with lower word-list delayed recall score (*B* = –5.59, 95% CI [ –10.31, –0.86], *P* = 0.021; Table 2). Removing the p-tau181 outlier and refitting the model produced a similar but stronger association (*B* = –7.58, 95% CI [–13.77, – 1.39], *P* = 0.017). No covariates were significantly associated with IC-CoDE memory domain.

Older age was associated with higher word-list delayed recall score in some models, namely the models that examined AT(N) profiles, p-tau181 (both with outlier included and removed), and p-tau217. No other covariates were associated with word-list delayed recall score.

## Discussion

This study provides comprehensive characterization of AT(N) blood biomarker profiles in older adults with epilepsy, demonstrating substantial heterogeneity in neurodegenerative signature. Only one-third of participants exhibited normal biomarker profiles, suggesting that neurodegenerative processes are common in epilepsy and may occur earlier or more frequently than expected based on age alone. Several important findings emerge from this work. First, abnormal biomarkers were associated with longer epilepsy duration and earlier seizure onset, suggesting that cumulative seizure exposure, epileptogenic activity, and chronic network dysfunction may contribute to AD-related pathological burden. Second, while categorical AT(N) classification was not associated with cognition, elevated p-tau181 was independently linked to poorer memory performance, suggesting that tau phosphorylation may be related to memory impairment in older adults with epilepsy. Third, biomarker selection substantially influenced both classification and clinical associations, underscoring important methodological considerations when applying the AT(N) framework to epilepsy.

### Distribution of AT(N) Profiles in Older Adults with Epilepsy

The distribution of AT(N) profiles in our epilepsy sample differed from that reported in healthy older adults. While studies of cognitively unimpaired individuals report normal biomarker profiles in 52-55% of participants, AD continuum profiles in 22-24%, and non-AD pathologic change in only 23-25% based on PET and/or CSF^39,40^, our sample showed a lower proportion with normal biomarkers (32%) and a higher proportion classified in the AD continuum (37%). These findings are broadly consistent with prior work demonstrating group-level age-accelerated alterations in amyloid and tau biomarkers in epilepsy, though results remain heterogeneous across cohorts depending on population characteristics and biomarker measurement methods^9,11,16,41^.

Our amyloid positivity rates appear broadly consistent with typical aging trends: 42% of the early-onset and 29% of the late-onset group were amyloid-positive, comparable to reported CSF positivity in adults of similar age without epilepsy or dementia^42^, and to plasma-based rates in the Alzheimer’s Disease Neuroimaging Initiative (ADNI) healthy control cohort^43^. However, the ADNI sample was older (mean age 72.6 years) and enriched for AD-related genetic risk (37.7% *APOE*-ε4 carriers), limiting direct comparison and suggesting our epilepsy sample likely carries elevated pathological burden for their age. Importantly, *APOE*-ε4 carrier status was higher in our AD continuum group, supporting that biomarker-based classification captures meaningful AD-related genetic risk rather than non-specific epilepsy-related processes alone.

### Demographic and Seizure Characteristics Associated with AD-related Pathology

Age and epilepsy duration were the only meaningful demographic differences across AT(N) profiles, and early-onset epilepsy was associated with higher odds of biomarker abnormality compared to LOUE despite being an average of 6.5 years younger. These results suggest that heightened biomarker abnormality in the early-onset group may stem from cumulative pathophysiology of repeated seizures, with long-term disease exposure playing a critical role in shaping neurodegenerative trajectories. Work demonstrating that childhood-onset epilepsy is associated with age-accelerated brain amyloid accumulation in late middle age supports this supposition^44^. Preclinical evidence further suggests these processes are mutually reinforcing: seizures promote amyloid-β release and aggregation while amyloid accumulation lowers seizure threshold^45^, and elevated tau burden increases seizure severity and frequency, creating a self-reinforcing cycle of neurodegeneration^9,24,25^.

### Cognition and AD-Related Pathology in Epilepsy

The 2024 AA diagnostic criteria define AD biologically, such that abnormal core biomarkers indicate AD-related pathophysiology even in the absence of a dementia syndrome^4^; however, the International Working Group recommends caution against equating biomarker positivity alone with a clinical diagnosis^46^. In older adults with epilepsy, we interpret biomarker abnormalities as evidence of co-occurring AD-related biological processes relevant to cognitive vulnerability, while recognizing they do not establish the etiology of cognitive symptoms in a given individual.

Consistent with this, blood-based AT(N) profiles were not significantly associated with cognitive outcomes, likely reflecting the multifactorial nature of cognitive impairment in epilepsy, including seizure burden, medication effects, and epilepsy-specific neuropathology independent of traditional AD biomarkers. This may be amplified in our heterogeneous sample including extratemporal cases where cognitive impairment may arise from non-AD mechanisms.

Additionally, a high proportion of participants had minimal or no cognitive impairment (67.9%), further limiting variability in outcomes. However, similar findings were reported in a pilot study comparing CSF AT(N) profiles in epilepsy, in which biomarkers did not differ between those with and without cognitive impairment in LOUE^17^. That said, when we examined individual biomarkers continuously, higher p-tau181 levels were independently associated with worse memory, suggesting early tau phosphorylation may have a more direct and domain-specific effect on memory-related processes, potentially related to the convergence of tau pathology and epilepsy-related injury within medial temporal lobe structures^23^. These results suggest that continuous biomarker measures may be more sensitive than categorical classifications for detecting clinically meaningful associations in epilepsy, particularly regarding cognition.

### Biomarker-Specific Considerations in Epilepsy

Participants with early-onset epilepsy demonstrated higher levels of p-tau217, NfL, and greater amyloid burden compared to LOUE, while p-tau181 did not differ between groups. The convergence of elevated p-tau217 and lower Aβ42/Aβ40 in the early-onset group is notable, as p-tau217 is tightly coupled to amyloid-related processes and has demonstrated superior diagnostic accuracy for AD pathology in head-to-head comparisons^32^. Yet when considered alongside the cognitive findings, an informative dissociation emerges; p-tau217 and amyloid burden differentiated onset groups, but neither was independently associated with cognitive performance, whereas p-tau181 was the only biomarker linked to memory outcome. Although speculative, this may reflect distinct biological roles along the AD pathological cascade. Plasma p-tau217 may be more sensitive to the presence and accumulation of upstream amyloid-driven pathology, whereas p-tau181 may more closely index downstream tau-mediated neuronal processes with a proximal effect on hippocampal-dependent memory function^47,48^. These findings raise the possibility that p-tau217 and p-tau181 capture partially distinct aspects of AD-related pathology in epilepsy, and that the optimal plasma tau marker for the AT(N) framework in this population remains unclear. Longitudinal studies with serial biomarker sampling will be needed to clarify whether their differential associations reflect genuinely distinct biological processes or differing sensitivity along a shared pathological cascade.

When hippocampal volume replaced NfL as the neurodegeneration marker, fewer participants were classified with non-AD pathologic change even with a liberal cutoff, and the proportion with normal biomarker profiles (48%) more closely approximated cognitively healthy older adults^39,40^, highlighting how biomarker choice heavily influences classification. NfL elevations may reflect seizure-related neuronal injury rather than AD-related neurodegeneration^36^, potentially overestimating non-AD pathologic change; this interpretation is supported by our finding that NfL was the only biomarker associated with epilepsy duration. Hippocampal volume, while more specific to localized and cumulative structural change, may underestimate neurodegeneration if atrophy has not yet manifested, is global in distribution, or if seizure-related hippocampal changes confound interpretation. Consistent with these trade-offs, hippocampal volume-based profiles were associated only with ASM count, while NfL-based profiles were associated with established neurodegeneration risk factors (age, epilepsy duration, *APOE* ε4), suggesting NfL captures broader clinically relevant variance^49^, albeit at the cost of reduced specificity to AD-related processes^4^. The absence of a significant correlation between the two markers further suggests they index partially distinct aspects of neurodegeneration. Together, these findings underscore the need for epilepsy-specific normative data and multimodal approaches to quantifying neurodegeneration in this population.

### Limitations and Future Directions

Several limitations warrant consideration. The modest sample size necessitated collapsing AT(N) profiles into three categories, precluding examination of the full eight-profile framework or subgroup interactions, and the predominance of cognitively intact participants likely limited variability in cognitive outcomes. Compared to CSF, plasma biomarkers have lower analyte concentrations, greater assay variability, and susceptibility to peripheral influences that may introduce misclassification. Specifically, a laboratory measure of kidney function (e.g., eGFR) was unavailable, and reliance on clinical CKD diagnosis may not fully account for subclinical renal impairment affecting biomarker clearance^38^. Additionally, while seizure-related neuronal injury may transiently elevate plasma biomarker levels, this was pertinent for only three participants who experienced a seizure within 24 hours of blood collection. Finally, plasma biomarker thresholds validated in typical aging populations may not translate directly to epilepsy, particularly for neurodegeneration markers sensitive to non-specific seizure-related neuronal injury.

Future work should integrate plasma biomarkers with multimodal neuroimaging and longitudinal cognitive assessments to validate classifications against established in vivo markers of both epilepsy and neurodegenerative pathology. The BrACE longitudinal design will enable characterization of biomarker and cognitive trajectories and help distinguish stable neurodegenerative processes from transient seizure-related fluctuations. Finally, inclusion of comparative cohorts, such as healthy older adults and individuals with mild cognitive impairment and AD without epilepsy, will enhance generalizability and help disentangle shared and disorder-specific mechanisms linking epilepsy and neurodegeneration.

### Conclusion and Clinical Implications

In this cohort of older adults with epilepsy, application of the AT(N) framework revealed substantial heterogeneity in AD-related biomarker profiles. Early-onset, chronic epilepsy was strongly associated with biomarker abnormalities, supporting a role for cumulative disease burden in shaping neurodegenerative risk. Although categorical AT(N) profiles were not associated with cognitive classification, continuous p-tau181 was related to memory performance. These findings also highlight important methodological considerations for applying the AT(N) framework in epilepsy, specifically that p-tau217 and p-tau181 may index partially distinct aspects of AD-related pathology, and that neurodegeneration markers may require multimodal approaches given the confounding influence of seizure-related injury on NfL. Collectively, this work suggests that current AD-centric frameworks may require adaptation for epilepsy populations.

Clinically, these findings underscore the potential utility of AD-related biomarkers for identifying individuals at risk for cognitive decline who may benefit from closer cognitive monitoring, particularly those with early-onset, chronic epilepsy. They also support coordinated management of epilepsy and cognitive comorbidities, including consideration of early referral to memory disorder clinics. As blood-based AD biomarkers gain regulatory approval, their application to epilepsy cohorts offers a promising pathway toward improved risk stratification and understanding of shared neurodegenerative processes. As disease-modifying therapies for AD advance, understanding how neurodegenerative processes intersect with epilepsy will be critical, particularly given the historical exclusion of patients with seizures from clinical trials^50^. Ongoing multimodal longitudinal work will determine whether AT(N) classifications predict distinct cognitive trajectories and elucidate the interacting effects of AD-related biomarkers and seizure activity over time.

## Supporting information

STROBE Checklist

## Acknowledgements

This research was supported by the National Institutes of Health (NIH; R01NS120976) and the Cleveland Clinic Epilepsy Center and Neurological Institute’s Center for Outcomes Research and Evaluation (NI-CORE). Given that this manuscript is the result of funding in whole or in part by the NIH, it is subject to the NIH Public Access Policy. Through acceptance of this federal funding, NIH has been given a right to make this manuscript publicly available in PubMed Central upon the Official Date of Publication, as defined by NIH.

