## Supplementary material for "AT(N) Framework in Older Adults with Epilepsy: Plasma Biomarkers and Associations with Demographic, Clinical, and Cognitive Features": STROBE Checklist

**STROBE Statement — Checklist for Cross-Sectional Studies**

| **Section/Topic** | **Item #** | **Checklist Item** | **Reported on page/section** |
| --- | --- | --- | --- |
| **TITLE AND ABSTRACT** | | | |
|  | 1a | Indicate the study's design with a commonly used term in the title or the abstract | Methods (Study Design) |
|  | 1b | Provide in the abstract an informative and balanced summary of what was done and what was found | Abstract |
| **INTRODUCTION** | | | |
| Background/Objectives | 2 | Explain the scientific background and rationale for the investigation being reported | Introduction |
|  | 3 | State specific objectives, including any pre-specified hypotheses | Introduction (final paragraph) |
| **METHODS** | | | |
| Study design | 4 | Present key elements of study design early in the paper | Methods (Study Design) |
| Setting | 5 | Describe the setting, locations, and relevant dates, including periods of recruitment, exposure, follow-up, and data collection | Methods (Study Design, Participants) |
| Participants | 6a | Cross-sectional study—Give the eligibility criteria, and the sources and methods of selection of participants | Methods (Participants) |
|  | 6b | Not applicable (no matching) | N/A |
| Variables | 7 | Clearly define all outcomes, exposures, predictors, potential confounders, and effect modifiers. Give diagnostic criteria, if applicable | Methods (Measures, Biomarkers and AT(N) profiles, Neuropsychological data) |
| Data sources/measurements* | 8 | For each variable of interest, give sources of data and details of methods of assessment (measurement). Describe comparability of assessment methods if there is more than one group | Methods (Biomarkers and AT(N) profiles, Neuropsychological data, Hippocampal volume) |
| Bias | 9 | Describe any efforts to address potential sources of bias | Methods (Statistical analyses – bootstrapped CIs, sensitivity analyses, outlier exclusion) |
| Study size | 10 | Explain how the study size was arrived at | Results (Study sample) |
| Quantitative variables | 11 | Explain how quantitative variables were handled in the analyses. If applicable, describe which groupings were chosen and why | Methods (Biomarkers and AT(N) profiles, Statistical analyses) |
| Statistical methods | 12a | Describe all statistical methods, including those used to control for confounding | Methods (Statistical analyses) |
|  | 12b | Describe any methods used to examine subgroups and interactions | Methods (Statistical analyses – early-onset vs. LOUE, continuous biomarker models) |
|  | 12c | Explain how missing data were addressed | Methods (Hippocampal volume); Results (Study sample) |
|  | 12d | Cross-sectional study—If applicable, describe analytical methods taking account of sampling strategy | Methods (Statistical analyses) |
|  | 12e | Describe any sensitivity analyses | Methods (Statistical analyses – bootstrapped CIs, ipsilateral HV, outlier exclusion) |
| **RESULTS** | | | |
| Participants | 13a | Report numbers of individuals at each stage of study—e.g. numbers potentially eligible, examined for eligibility, confirmed eligible, included in the study, completing follow-up, and analysed | Results (Study sample) |
|  | 13b | Give reasons for non-participation at each stage | Results (Study sample – excluded due to missing AT(N) data) |
|  | 13c | Consider use of a flow diagram | Not included; exclusion described in text |
| Descriptive data | 14a | Give characteristics of study participants (e.g. demographic, clinical, social) and information on exposures and potential confounders | Results (Study sample); Table 1 |
|  | 14b | Indicate number of participants with missing data for each variable of interest | Methods (Hippocampal volume); Results (Study sample) |
|  | 14c | Not applicable (cohort-specific item) | N/A |
| Outcome data* | 15 | Cross-sectional study—Report numbers of outcome events or summary measures | Results (AT(N) profile distribution; cognitive outcomes) |
| Main results | 16a | Give unadjusted estimates and, if applicable, confounder-adjusted estimates and their precision (e.g. 95% CI). Make clear which confounders were adjusted for and why they were included | Results (Associations between AT(N) profiles and cognition; continuous biomarker analyses) |
|  | 16b | Report category boundaries when continuous variables were categorized | Methods (Biomarkers and AT(N) profiles – cutpoints reported explicitly) |
|  | 16c | If relevant, consider translating estimates of relative risk into absolute risk for a meaningful time period | N/A (cross-sectional) |
|  | 16d | Report results of any adjustments for multiple comparisons | Methods/Results (no formal correction applied; p<0.05 threshold stated) |
| Other analyses | 17a | Report other analyses done—e.g. analyses of subgroups and interactions, and sensitivity analyses | Results (Secondary analyses: continuous biomarkers, p-tau217, hippocampal volume substitution) |
|  | 17b | If numerous genetic exposures were examined, summarize results from all analyses undertaken | Results (APOE-ε4 reported as covariate/group characteristic) |
|  | 17c | If detailed results are available elsewhere, state how they can be accessed | N/A |
| **DISCUSSION** | | | |
| Key results | 18 | Summarise key results with reference to study objectives | Discussion (opening paragraph) |
| Limitations | 19 | Discuss limitations of the study, taking into account sources of potential bias or imprecision. Discuss both direction and magnitude of any potential bias | Discussion (Limitations) |
| Interpretation | 20 | Give a cautious overall interpretation of results considering objectives, limitations, multiplicity of analyses, results from similar studies, and other relevant evidence | Discussion |
| Generalisability | 21 | Discuss the generalisability (external validity) of the study results | Discussion (Limitations – single-site recruitment, predominantly white/female sample) |
| **FUNDING** | | | |
|  | 22 | Give the source of funding and the role of the funders for the present study and, if applicable, for the original study on which the present article is based | Funding section |

** Give information separately for cases and controls in case-control studies and, if applicable, for exposed and unexposed groups in cohort and cross-sectional studies.*
